# Quality testing of mifepristone and misoprostol in 11 countries

**DOI:** 10.1101/2023.07.10.23292436

**Authors:** Jason Bower, Lester Chinery, Alessandra Fleurent, A. Metin Gülmezoglu, Wallada Im-Amornphong, Catherine Kilfedder, Petra Procter, Alessandra Tomazzini

**Affiliations:** Independent consultant, London, UK; Concept Foundation, Geneva, Switzerland; International Planned Parenthood Federation, London, UK

**Keywords:** Quality, Abortion, Misoprostol, Mifepristone

## Abstract

**Objective:** Previous studies have demonstrated quality concerns with misoprostol. Mifepristone, however, has not been extensively assessed for quality. Between 2020 and 2021, Concept Foundation and the International Planned Parenthood Federation conducted a study to determine the quality of these medical abortion drugs in low– and middle-income countries (LMIC).

**Methods:** Collection of batch samples of misoprostol and mifepristone was carried out by trained sampling agents in selected LMIC. Single drug packs and combipacks were sampled. A World Health Organization prequalified laboratory conducted testing method verifications and subsequent sample analysis. Tests included identification, assay, related substances, and content uniformity for misoprostol, and identification, assay, related substances, and dissolution for mifepristone.

**Results:** Samples were collected from Burkina Faso, Cambodia, Democratic Republic of Congo, India, Kyrgyzstan, Moldova, Nepal, Nigeria, Pakistan, Uganda and Vietnam. Sixty-four pooled batch samples were tested, consisting of 31 combipacks, 26 misoprostol-only and 7 mifepristone-only products. Overall, 54.7% of samples were non-compliant with one or more of the specifications, representing 51.6% of combipack products, 57.1% of misoprostol tablets analyzed and 23.7% of mifepristone tablets. One falsified misoprostol-only product was found.

**Conclusion:** This study confirms that a significant problem still exists in relation to the quality of medical abortion drugs in low– and middle-income countries. For misoprostol, our findings suggest that historical concerns around primary packaging may have been largely resolved but that manufacturing processes for both finished product and active pharmaceutical ingredient need to be improved. This study also provides evidence of mifepristone quality issues.

## Background

Three out of ten pregnancies worldwide end in induced abortion, with medical methods playing an increasingly important role [1]. Medical abortion is usually conducted through the administration of either a combination of mifepristone, a synthetic antiprogesterone, and misoprostol, a prostaglandin analogue, or misoprostol alone [2]. Both drugs are included on the World Health Organization (WHO) Essential Medicines List (EML) for medical abortion. Misoprostol is also recommended for cervical ripening, labor induction, and prevention and treatment of postpartum hemorrhage [3]. Medical abortion provides individuals who are pregnant with a non-invasive alternative to surgical abortion and reduces the need for skilled surgical abortion providers, which is particularly helpful in low-resource environments [2,4,5]. Ensuring that quality misoprostol and mifepristone are accessible worldwide is crucial to reduce the number of unsafe abortions, which make up about 45% of all abortions [1]. The majority of unsafe abortions take place in low-and middle-income countries (LMIC).

There are limited options for the procurement of quality-assured misoprostol and mifepristone, with very few products demonstrably meeting the quality standards defined by WHO’s Prequalification of Medicines Program [6] or Stringent Regulatory Authorities (SRA) (as defined in the WHO Technical Report Series no. 1003 [7]), the minimum standard widely adopted by United Nations (UN) agencies, United States Agency for International Development (USAID) and other major donors [8,9]. Furthermore, the limited number of quality-assured medical abortion drugs are not widely procured and distributed in LMIC, likely because of purchase prices that are considered too high within the existing supply chain mechanisms. Consequently, women and girls in LMIC are at risk of utilizing medical abortion drugs that have not undergone stringent quality assessments for safety and efficacy.

Use of substandard and falsified medicines can have a severe impact on public health and are especially prevalent in LMIC [10,11]. Poor quality medical abortion drugs, such as those with less active pharmaceutical ingredient (API) than needed, can result in failed or incomplete abortion, and may require further medical management [2].

Numerous studies conducted to examine the quality of misoprostol in circulation at country level in LMIC have indicated that drug quality is sub-optimal [12–15]. Misoprostol is a viscous oil at room temperature and is extremely unstable in the presence of moisture; therefore, both the API and finished pharmaceutical product (FPP) tablets need to be manufactured under stringently controlled conditions and be packaged appropriately in double-sided aluminum (Alu/Alu) blisters [12,13,16].

Mifepristone quality in LMIC has not been extensively studied, as it is a more stable molecule than misoprostol.

The objective of this study was to assess whether the quality of misoprostol has improved over the years and explore whether quality issues may also be present for mifepristone to identify key areas of medical abortion drug quality that can be addressed by future interventions.

## Methods

Countries were selected based on their LMIC status in 2020, market size, availability of sampling partners on the ground, and variety of medical abortion drugs in each market [17]. Countries with local manufacturing of medical abortion drugs were prioritized as these products are often exported to other LMIC. Samples were collected from: Bangladesh, Burkina Faso, Cambodia, Democratic Republic of Congo, India, Kyrgyzstan, Moldova, Nepal, Nigeria, Pakistan, Uganda and Vietnam. Convenience sampling was employed as the study was carried out independently of local regulatory authorities. Standard Operating Procedures (SOPs) and a Sample Collection Form (SCF) were developed and shared with focal points in each country who were familiar with their national medical abortion markets. Sampling agents were trained on the SOPs and then collected samples of products of misoprostol (200 mcg) tablets, mifepristone (200 mg) tablets, and the mifepristone/misoprostol tablets “combipack”, containing one mifepristone 200 mg tablet and four misoprostol 200 mcg tablets. Agents used either the Mystery Shopper approach, Overt Sampling, or a hybrid of the two methods. The target minimum sample size of each type of pooled batch sample was: 35 misoprostol tablets, 15 mifepristone tablets, and 15 combipacks. Agents collected batches that were more than six months beyond their date of manufacture, to provide a better indication of the product quality throughout its shelf-life. Sites were primarily at the client point-of-purchase or service provision including pharmacies, hospitals and drug sellers.

Data on each batch sample was validated in-country and centrally. The sampling agents then shipped the samples via reputable international courier companies to the testing laboratory in Germany, where samples were kept in a temperature-controlled room between 15°C and 25°C. The sampling agents in Bangladesh were unable to ship the samples for testing.

Given the anticipated difficulty in sampling these restricted products, and to optimize the number of samples that could be tested, sample quality analysis was limited to testing parameters considered most critical to indicate clinically relevant quality issues in misoprostol and mifepristone tablets.

For misoprostol, identification, assay and related substances were considered essential, given the link to efficacy and the known instability of the product. Due to the low ratio of API content to tablet mass and therefore higher risk of non-homogeneity within batches, content uniformity was included. Dissolution or disintegration were not included as misoprostol is a highly soluble substance [18]. Misoprostol tablets were analyzed for the selected parameters using the methods and specifications described in the International Pharmacopoeia (Ph. Int.), Misoprostol tablets monograph (2016-01) [19].

For mifepristone, the parameters selected were identification, assay, related substances, and dissolution. Mifepristone is not a highly soluble substance [20], and hence may pose a higher risk of poor tablet dissolution, affecting product bioavailability. Uniformity of mass was not tested due to the large API component in tablets and hence low risk of non-homogeneity in production.

Currently, no published international reference pharmacopeial methods and specifications exist for mifepristone API or FPP. In-house methods for assay, related substances determination and dissolution were therefore developed and validated. The specifications were established based on a 2008 United States Pharmacopeia (USP) pending monograph for mifepristone API [21], the International Council for Harmonization of Technical Requirements for Pharmaceuticals for Human Use (ICH) Q3B guideline [22], as well as the analytical results of the innovator product, Mifegyne (Nordic Pharma). Details of the specifications of misoprostol and mifepristone tablets used in the study are shown in Tables S1 and S2, Additional file 1.

Analytical method verification and sample analysis was carried out by the WHO-prequalified quality control laboratory, Institute for Pharmaceutical and Applied Analytics (InphA), Bremen, Germany, as outlined in Table S3, Additional file 1.

The samples to be investigated were adjudged in compliance, if they met the specifications outlined in Tables S1 and S2, Additional file 1. For overall results, a sample was rated overall as non-compliant if it failed any of the tests conducted.

## Results

Sampling was carried out between December 2020 and September 2021, at 50 sites in 12 countries, across five of the six regions defined by WHO [23]. Since the nine samples collected in Bangladesh were not tested, the analysis is based on samples from 11 countries. Data was recorded for 84 different batch samples in total; of these, 64 were tested (Figure 1). Further collection data are presented in Tables S4 and S5, Additional file 1.

**Figure 1:**
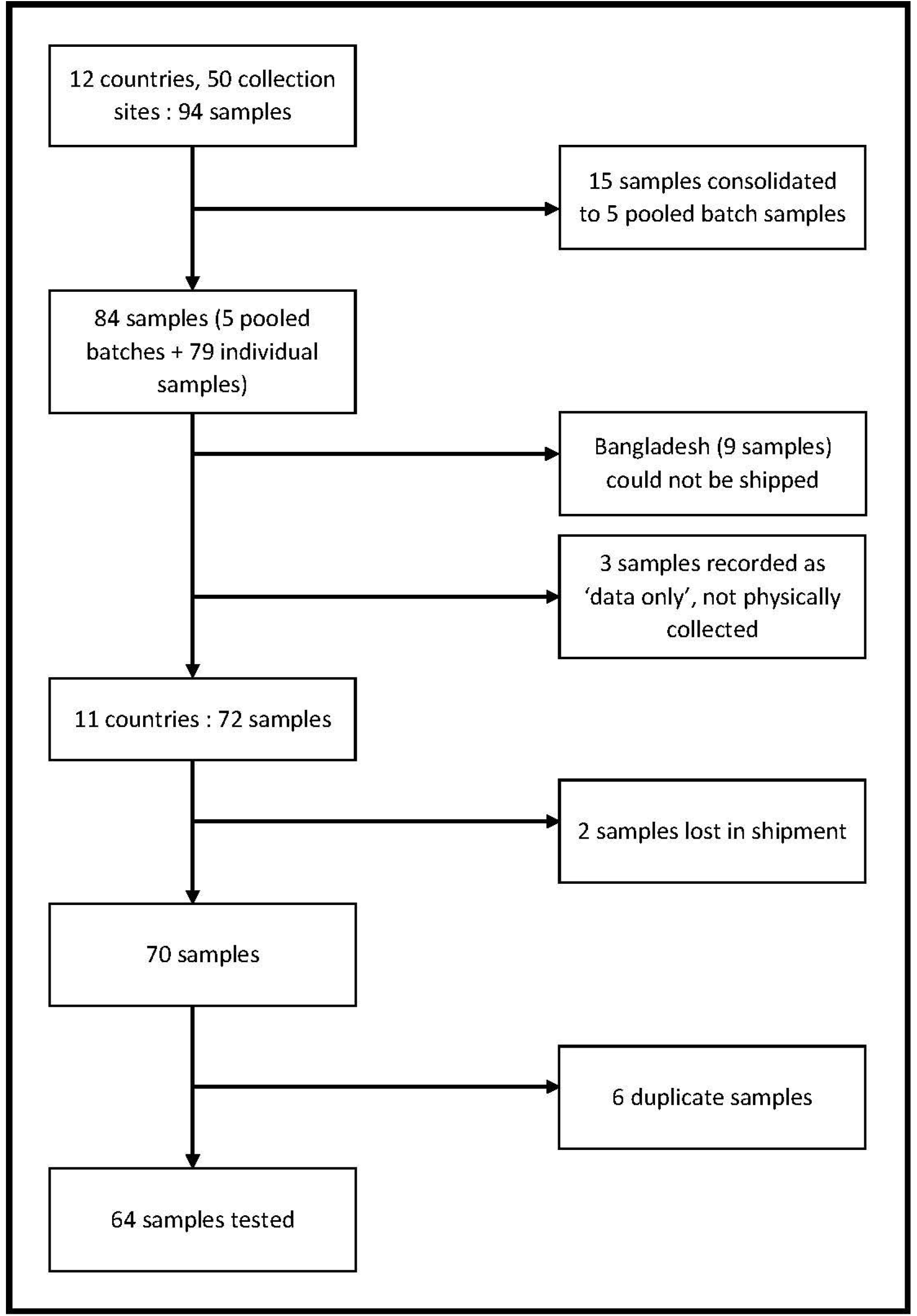
Sample collection and analysis flow diagram.

Purchase prices were recorded for 91 samples. The unit sizes used were: 1 pack for combipack products; or 1 tablet for misoprostol-only and mifepristone-only products. Unit prices of each sample were converted from local currency into USD, using the online currency converter https://www.xe.com/currencytables/. The median price for combipack samples was $5.70 per pack (range $2.49 – $14.10; n=50). The median unit prices for misoprostol-only products were $0.43 per tablet ($0.08 – $1.75; n=33), and $3.90 per tablet ($1.97 – $17.36; n=8) for mifepristone-only products.

The 64 samples tested consisted of 31 combipack products, 26 misoprostol-only products, and 7 mifepristone-only products (Table 1). Therefore, 57 samples contained misoprostol 200 mcg tablets and 38 contained mifepristone 200 mg tablets (Figure 2).

**Figure 2:**
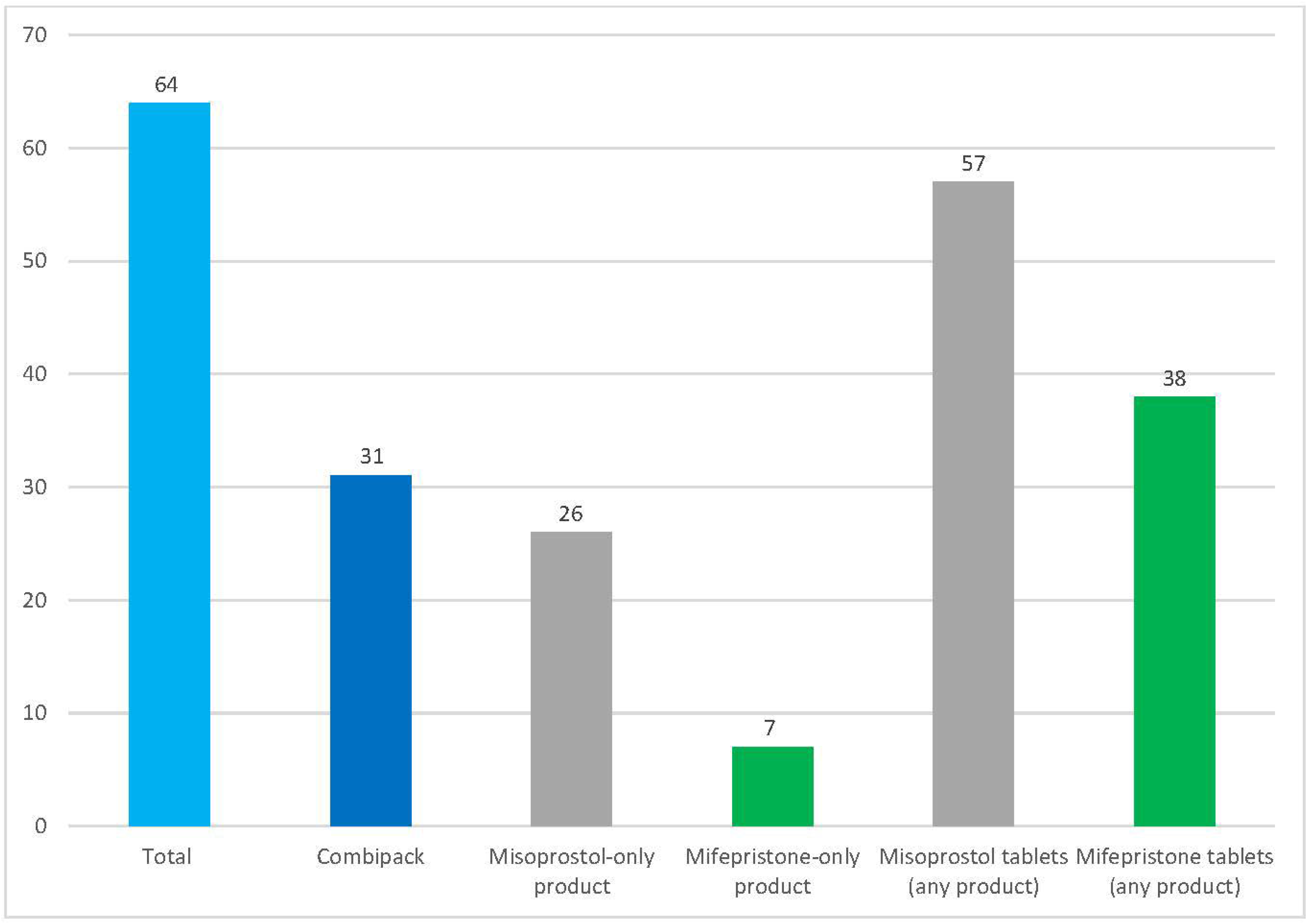
Samples tested per product type and active ingredient type.

**Table 1:**
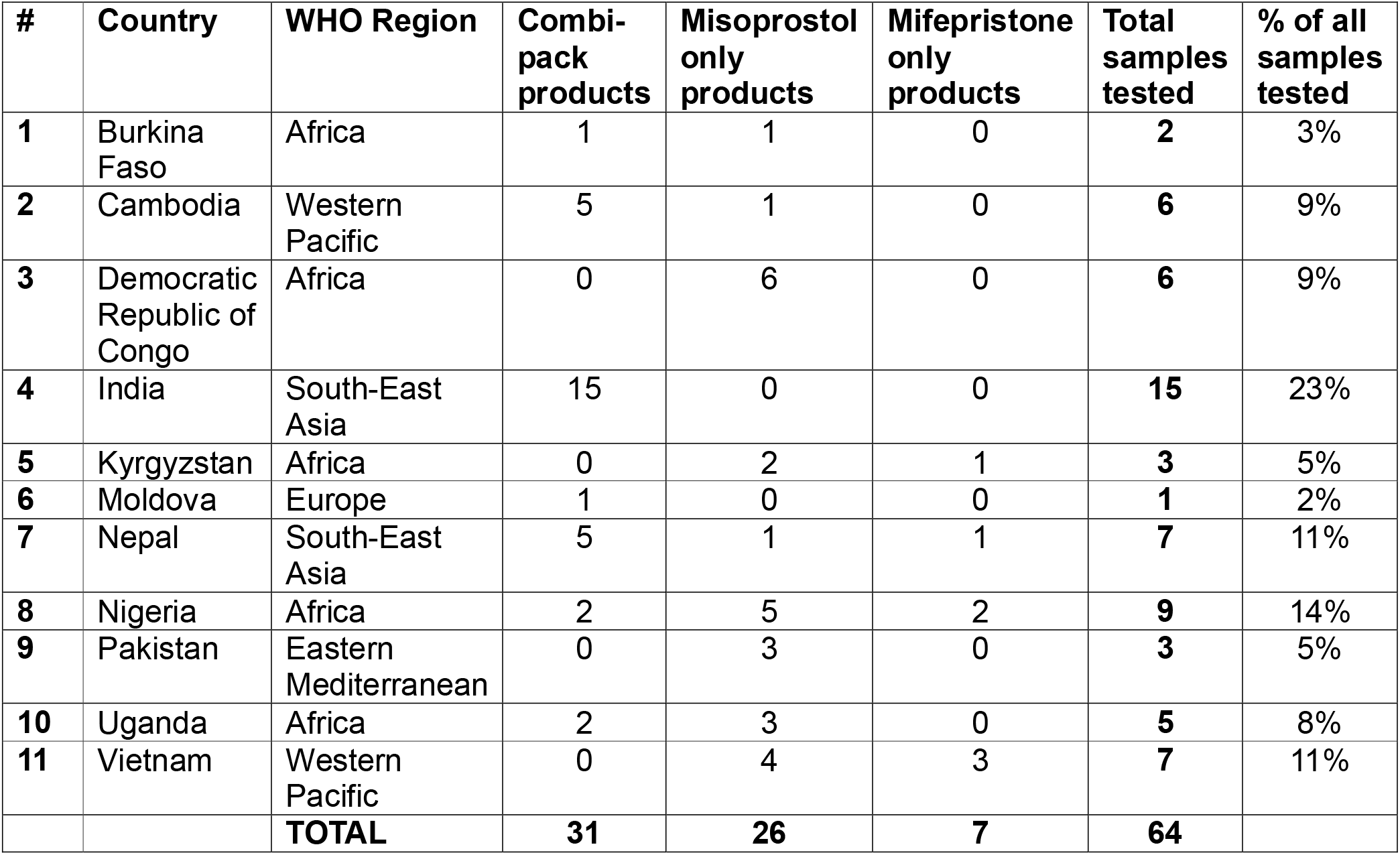
Samples tested per country

| # | Country | WHO Region | Combi-pack products | Misoprostol only products | Mifepristone only products | Total samples tested | % of all samples tested |
| --- | --- | --- | --- | --- | --- | --- | --- |
| 1 | Burkina Faso | Africa | 1 | 1 | 0 | 2 | 3% |
| 2 | Cambodia | Western Pacific | 5 | 1 | 0 | 6 | 9% |
| 3 | Democratic Republic of Congo | Africa | 0 | 6 | 0 | 6 | 9% |
| 4 | India | South-East Asia | 15 | 0 | 0 | 15 | 23% |
| 5 | Kyrgyzstan | Africa | 0 | 2 | 1 | 3 | 5% |
| 6 | Moldova | Europe | 1 | 0 | 0 | 1 | 2% |
| 7 | Nepal | South-East Asia | 5 | 1 | 1 | 7 | 11% |
| 8 | Nigeria | Africa | 2 | 5 | 2 | 9 | 14% |
| 9 | Pakistan | Eastern Mediterranean | 0 | 3 | 0 | 3 | 5% |
| 10 | Uganda | Africa | 2 | 3 | 0 | 5 | 8% |
| 11 | Vietnam | Western Pacific | 0 | 4 | 3 | 7 | 11% |
|  |  | **TOTAL** | **31** | **26** | **7** | **64** |  |

The 64 tested samples were manufactured by 35 distinct manufacturers, from nine different countries of origin. 12 products tested were either approved by an SRA as defined by WHO (as defined in the WHO Technical Report Series no. 1003 [7]), or prequalified by the WHO Prequalification Program (PQP) [6].

15 (of the 64) samples were products marketed by international social marketing organizations (SMO), including eight marketed for individual domestic markets and seven exported to international markets.

The shelf-life of products ranged from 12 months to 48 months, as shown in Table S6, Additional file 1. All samples tested were beyond six months of their date of manufacture and were within their shelf-life at the time of testing.

Sample analysis was performed at the laboratory between February and November 2021.

Most samples were of sufficient quantity for the critical test parameters listed in Tables S1 and S2, Additional file 1. Exceptions were three combipack samples: for two, all tests except mifepristone dissolution were conducted; for another, mifepristone dissolution was the only test conducted. As a result, out of 38 total tested mifepristone samples, two were tested for assay and related substances only, one was tested for dissolution only, and 35 were tested for all parameters. For misoprostol, 56 out of 57 samples were tested, but one misoprostol-only product failed the identification test, so further tests on that sample were not conducted. Therefore, a total of 55 samples were tested for all other misoprostol test parameters.

Of 64 samples tested, 35 (54.7%) were found to be OOS for at least one test parameter and reported as non-compliant with the established specifications (Figure 3). 16 out of 31 combipacks (51.6%) were found non-compliant, as were 16 of 26 misoprostol-only products (61.5%), and three of seven mifepristone-only products (42.9%). For the 31 combipack samples tested, both the misoprostol tablet and the mifepristone tablet were each OOS in six (19.4%) samples, whereas only the misoprostol tablet was OOS in a further ten (32.3%) samples.

**Figure 3:**
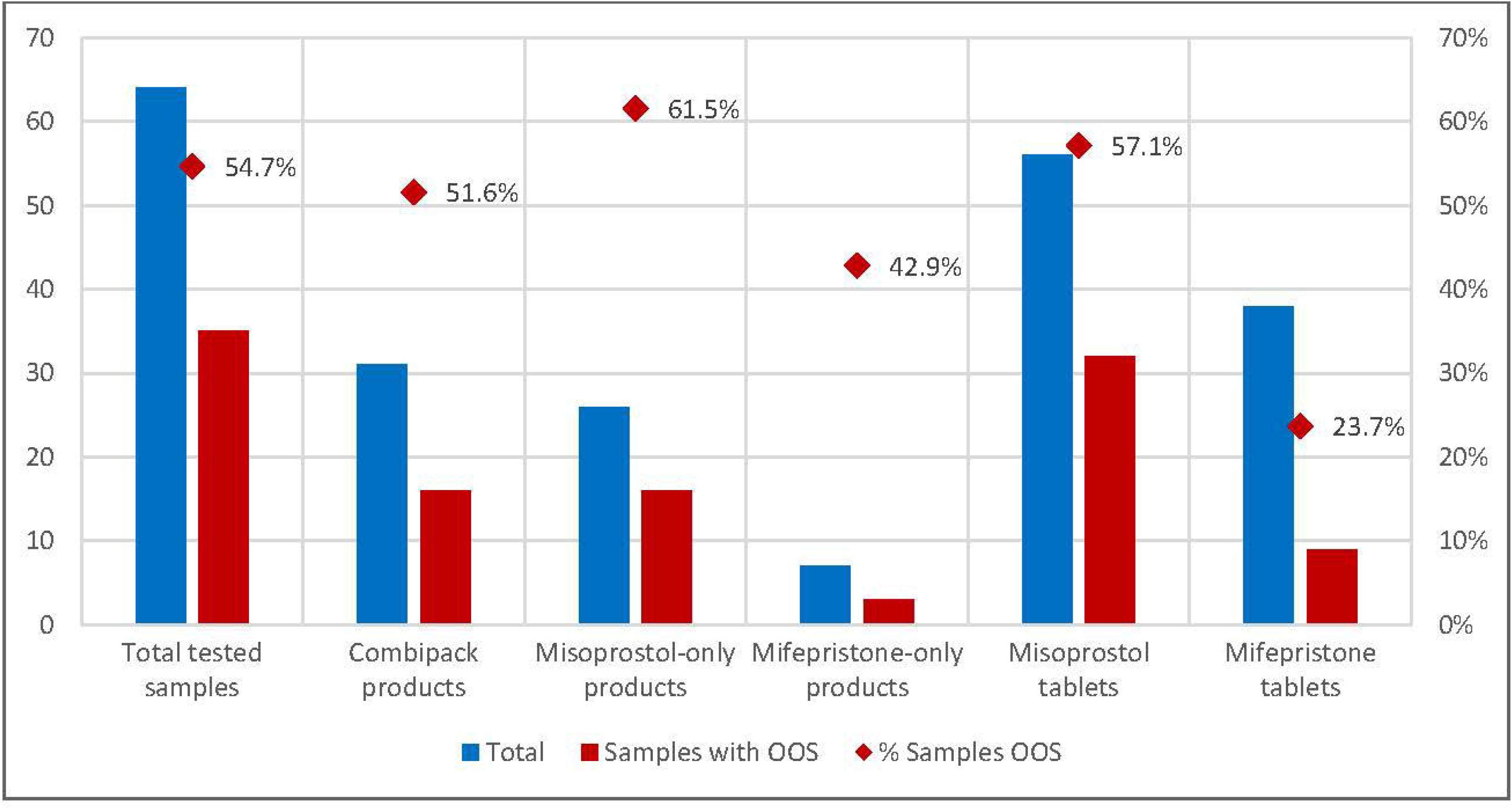
Non-compliant findings per product type and active ingredient type.

Out of 56 misoprostol tablet samples tested, 32 (57.1%) were observed to be non-compliant. Of 38 mifepristone tablet samples tested, nine (23.7%) were found to be non-compliant with specifications. See Table S7 and S8, Additional file 1, for summaries of misoprostol and mifepristone samples testing results respectively.

### Misoprostol tablet findings

For misoprostol tablets, the highest proportion of non-compliant findings were observed for related substances Impurity C, followed by Sum of Impurities A, B and E (Figure 4).

**Figure 4:**
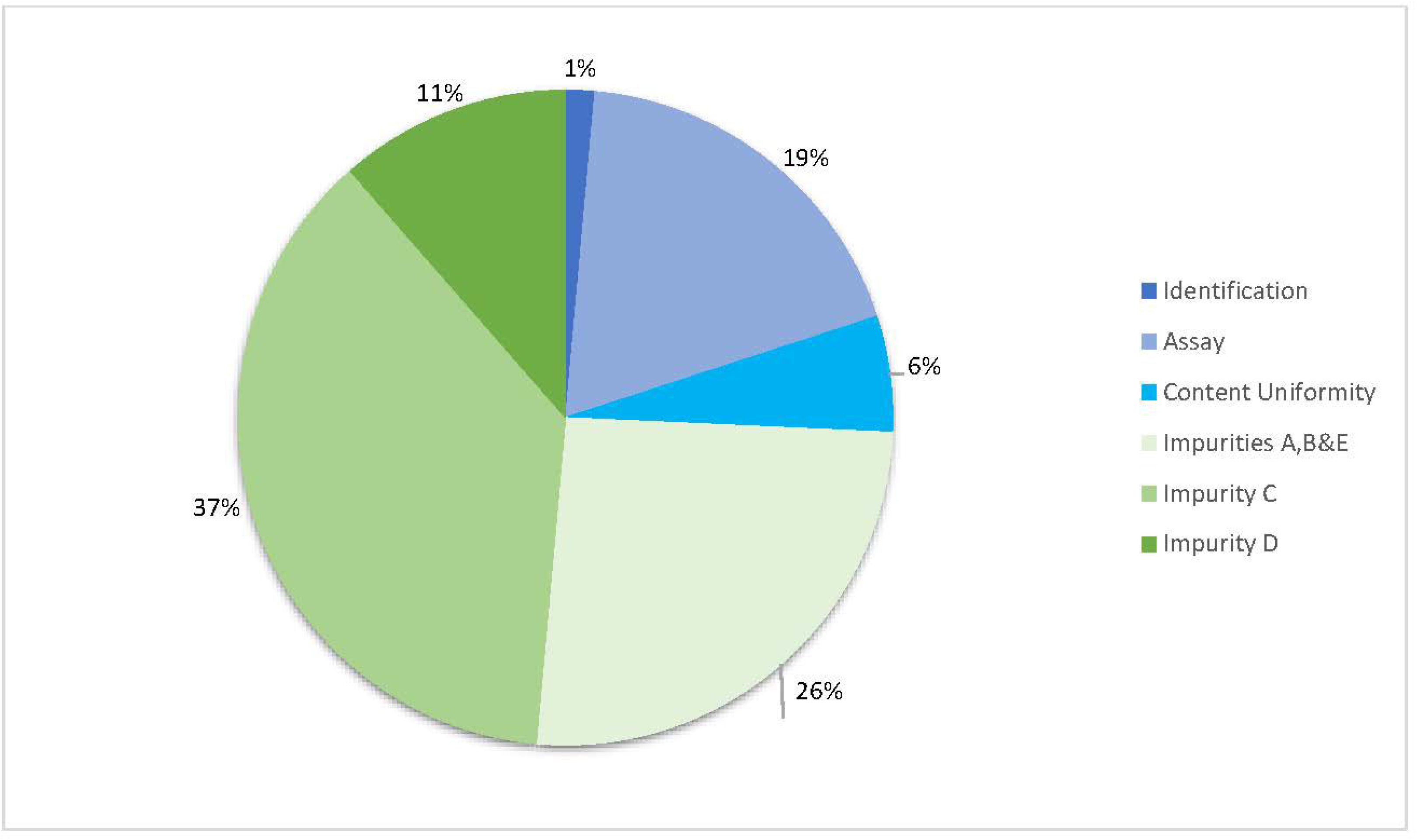
OOS profile for misoprostol tablets (proportion out of n=70 total non-compliant findings).

Misoprostol identification and assay was found to be OOS in 1 of 56 (1.8%) and 13 of 55 (23.6%) samples respectively. Content uniformity was OOS in the four samples with the lowest assay values (13.4-75.3%) but was within specification for other samples. Any related substance criteria were OOS in 31 of 55 samples tested (56.4%). The condensed scale of related substances findings is shown in Figure 5, whereas the full scale of this figure is provided as Figure S1, Additional file 2.

**Figure 5:**
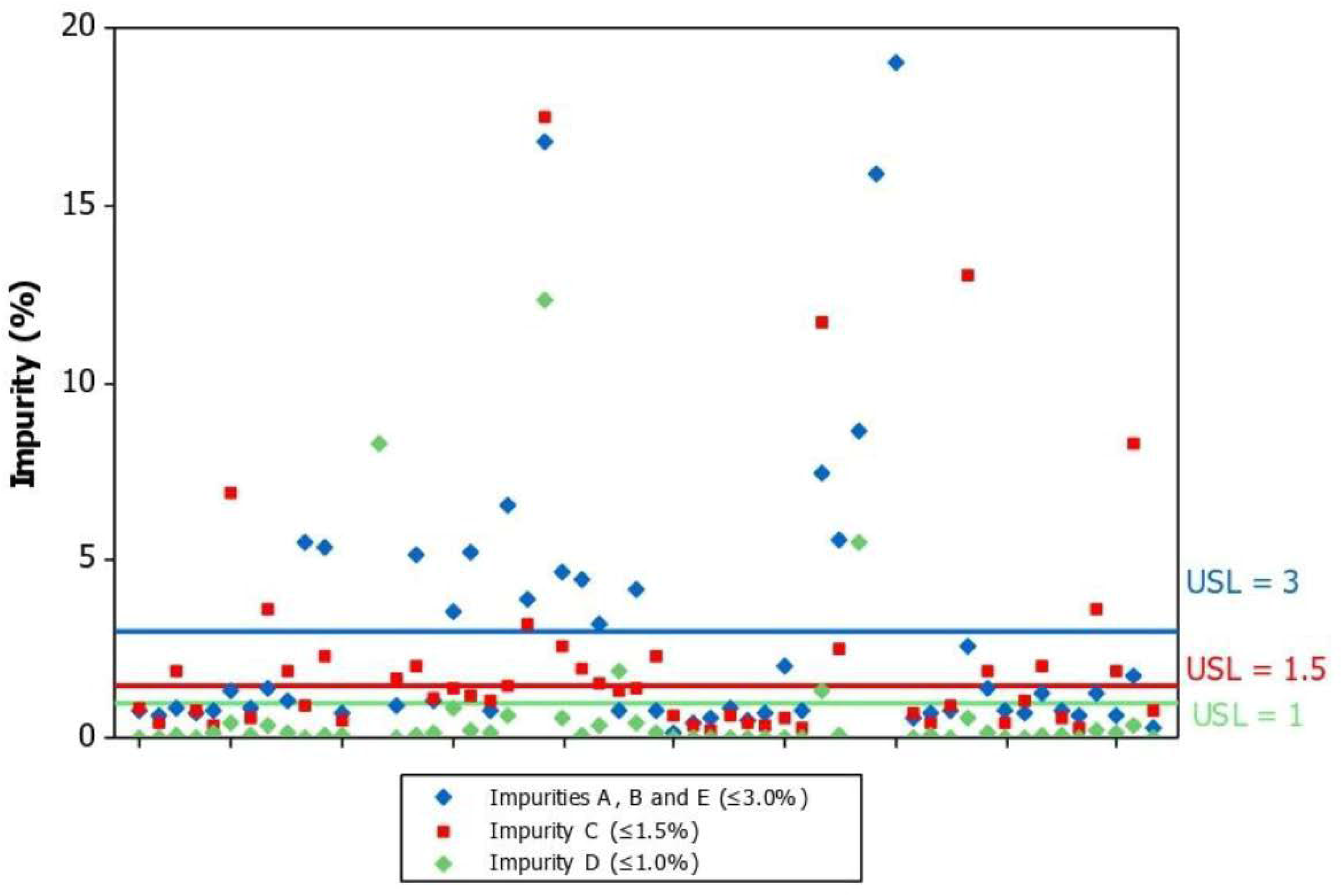
Misoprostol related substances results (condensed scale).

**Figure 6:**
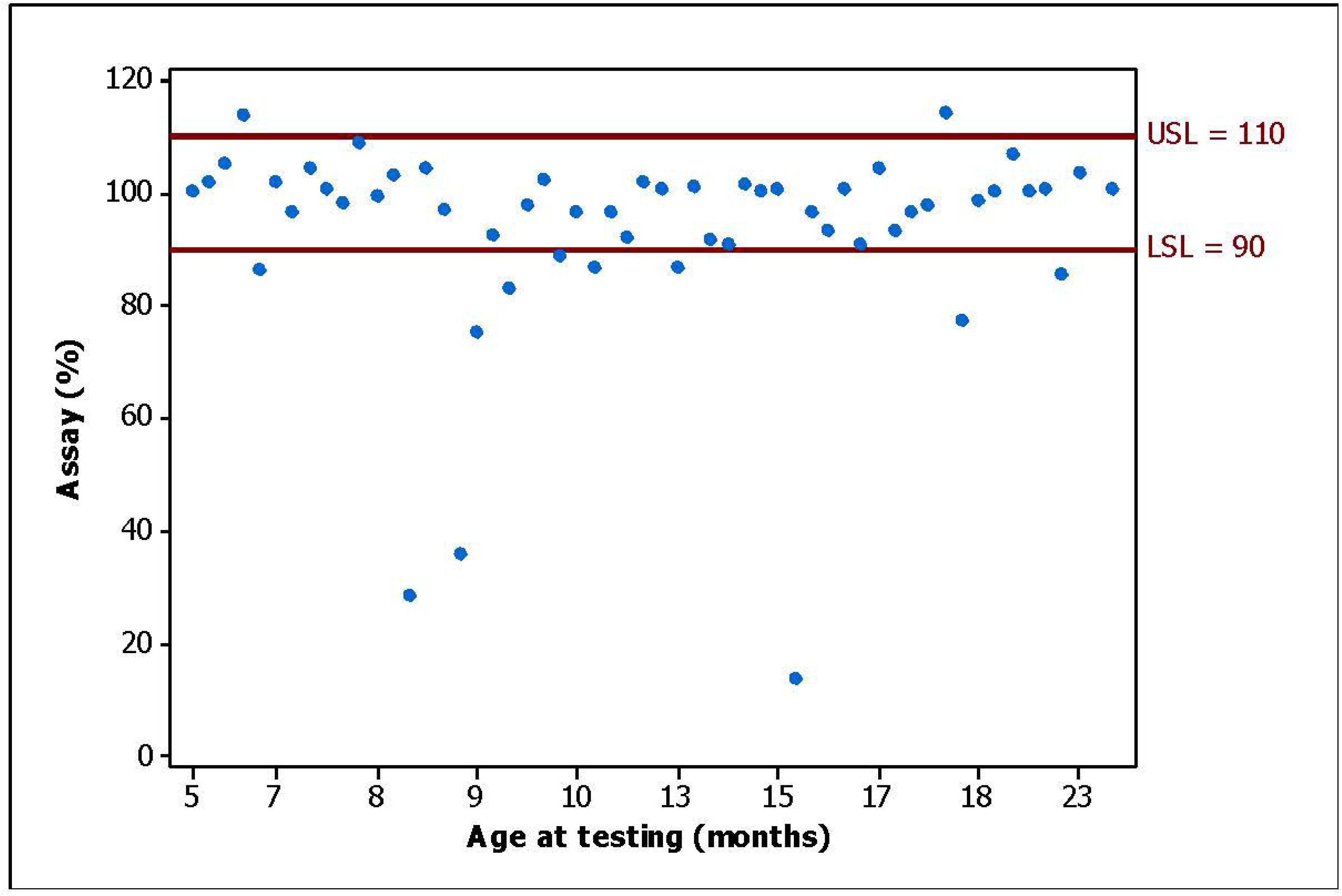
Misoprostol assay results per age of sample.

Excluding the sample that failed the identification test with no misoprostol content, the three worst misoprostol assay results observed were 13.4%, 28.4%, and 35.7%. A further eight samples ranged between 75-90%.

Misoprostol assay was assessed against the age of the sample when tested. As seen in Figure 6, there was no correlation observed.

Misoprostol assay versus type of primary packaging was not examined further, as 55 of the 56-misoprostol tablet-containing products were packaged with aluminum foil on both sides (Alu/Alu blister).

Mood’s Median Test was conducted to assess correlation between observed storage conditions at site and misoprostol assay, and labelled storage requirements and misoprostol assay. No statistically significant difference was found for either test.

Finally, the Mood’s test and box chart analysis also showed no significant differences between the median purchase price of compliant products versus those of OOS products.

### Falsified Cytotec®

One Cytotec (Piramal Healthcare Ltd, United Kingdom) sample, collected from a pharmaceutical wholesaler, failed the initial misoprostol identification test. Irregularities were noted on tablet disintegration during sample solution preparation.

Differences in tablet color and package insert were noted between this sample and an otherwise identical Cytotec product from a nearby country (see Figure 7). Retesting was carried out which confirmed the negative identification result. Further analysis to screen for undeclared active ingredients was performed by HPLC/mass spectrometry and was also negative.

**Figure 7:**
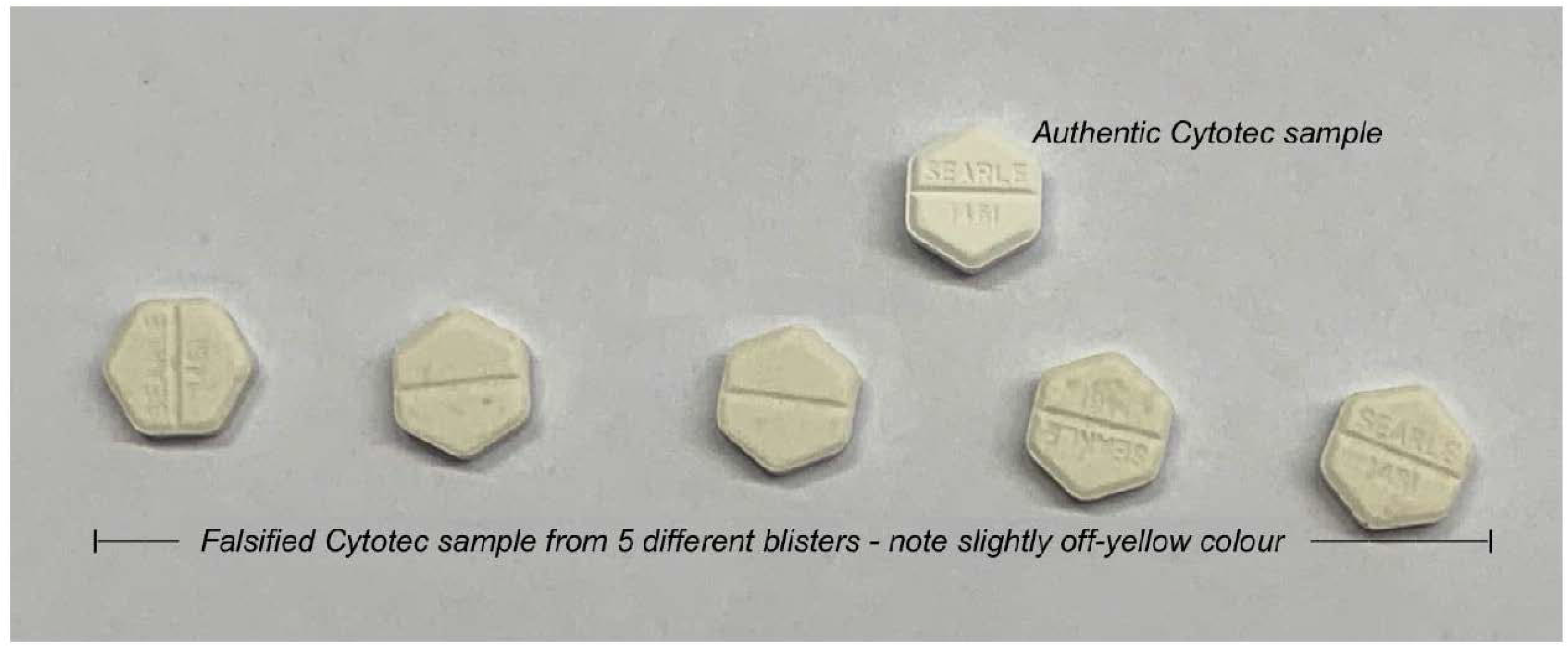
Differences in tablet appearance between authentic product and falsified product.

A report was submitted to the WHO Incidents and Substandard/Falsified Medical Products (ISF) team for further investigation, which later revealed the presence of the same falsified batch in two neighboring countries, and a second falsified batch of the same product in two neighboring countries. Subsequently, WHO issued a falsified product alert [24].

### Mifepristone tablet findings

For mifepristone tablets, the highest proportion of non-compliant findings was observed for related substances, specifically any individual unknown impurity (Figure 8). No OOS result was found for the demethylated derivative impurity of mifepristone (Figure 9).

**Figure 8:**
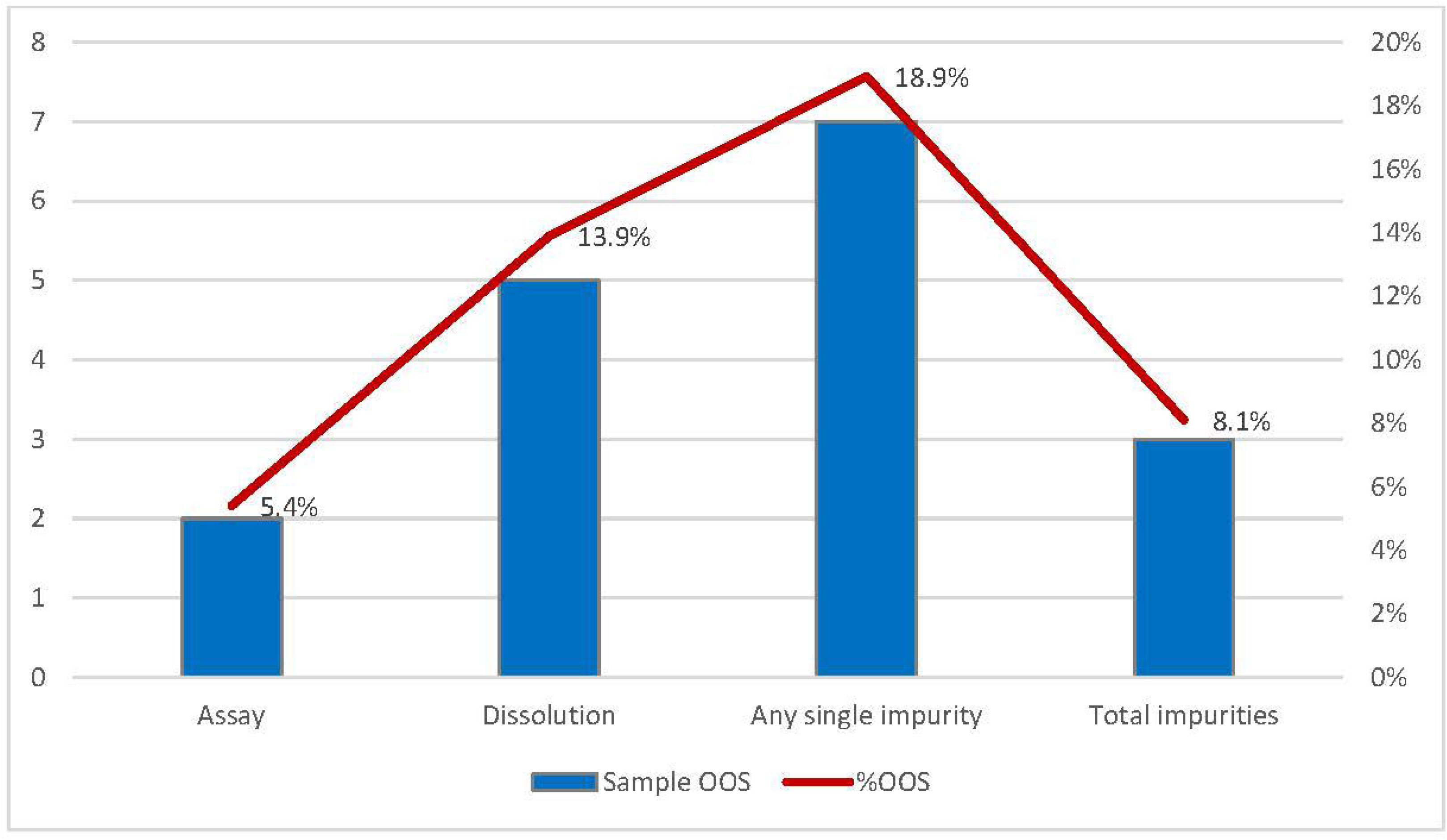
Rate of OOS per mifepristone tablet test parameter.

**Figure 9:**
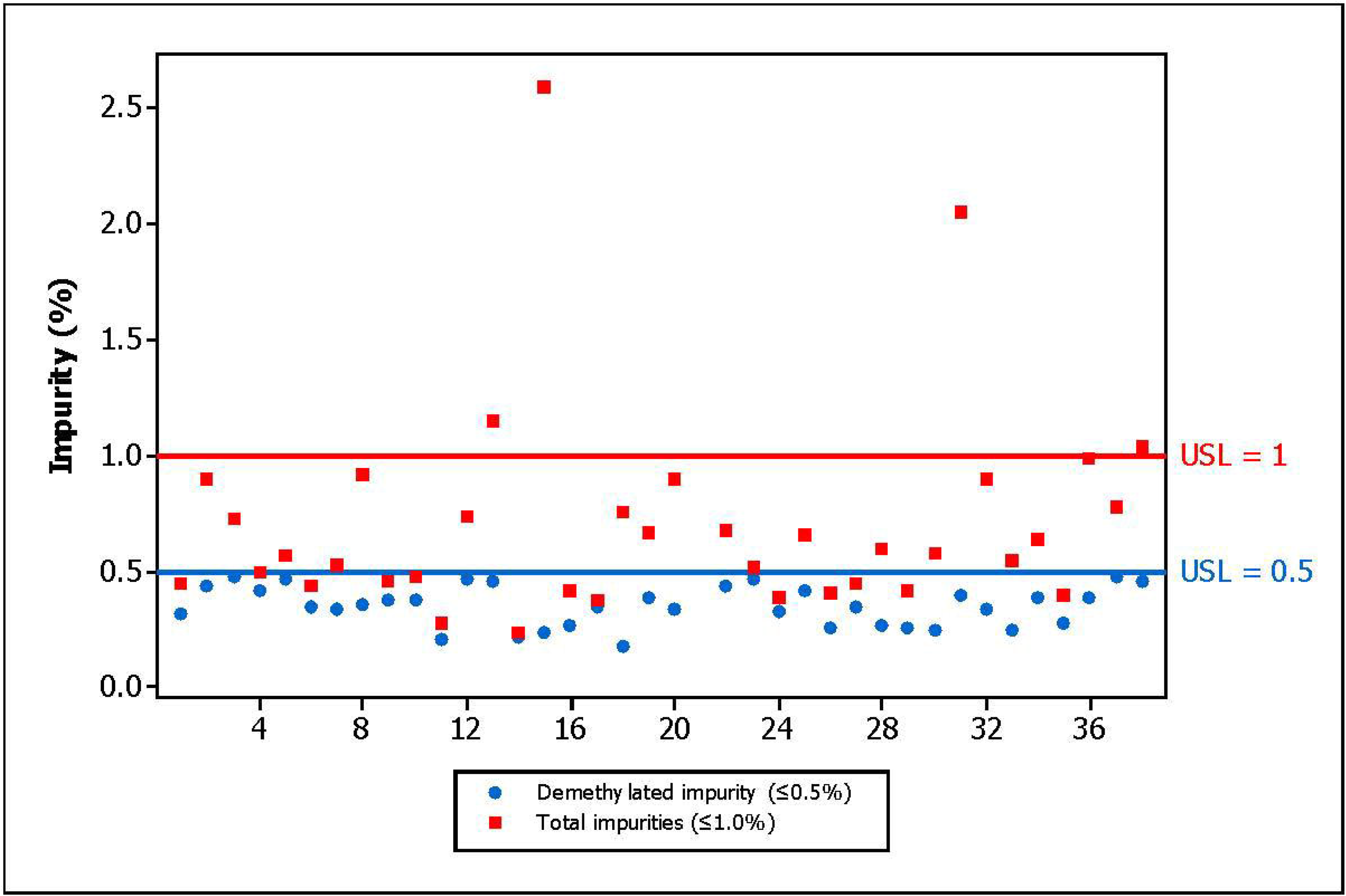
Mifepristone related substances results for each sample.

Mifepristone assay was found to be OOS in two of 37 (5.4%) samples, measuring 78.6% and 84.5% content (see Figure S2, Additional file 3). Related substances overall were found to be OOS in eight of 37 (21.6%) samples. Five samples of mifepristone (13.9%) were observed to fail stage 1 dissolution tests. Of these, two samples were major OOS (averaging 11% and 53% content dissolved in 30 minutes), and three samples were minor OOS whereby the stage 2 testing was warranted but not possible due to insufficient samples.

### Other variables

Products were manufactured in nine different countries, with most samples originating from India (67.2%). Products manufactured in India were distributed both locally and internationally. Products manufactured in China, Republic of Korea, Russia, and the United Kingdom (UK) were all exported, whereas products manufactured in other countries were sampled in their local market.

26 out of 43 (60%) Indian-manufactured samples were found to be OOS. High rates of non-compliance were also seen in the samples manufactured in Vietnam (four out of six) and Pakistan (two out of three). No OOS findings were observed in the small number of samples manufactured in China, Nepal, Nigeria and the UK.

Excluding the falsified sample, of 11 remaining samples tested holding SRA approval or WHO Prequalification status, one misoprostol-only sample was non-compliant. It was noted that this non-compliant sample was OOS on Impurity C only (result 1.9%, acceptable limit ≤1.5%); the assay result was 100.6% and the sample had aged 22 months of its 24-month shelf-life when tested. Of the 52 samples tested that do not hold SRA approval or WHO Prequalification status, 33 (63.5%) were non-compliant.

Of the 15 products marketed by SMOs, four (25%) were non-compliant with specifications, compared to 63.3% for the remaining products. All were domestically distributed combipack products (see Figure S3, Additional file 4).

## Discussion

We found that 54.7% of medical abortion samples from 11 LMIC were non-compliant with the specifications applied. Combipack products were non-compliant in 51.6% of samples, whereas misoprostol tablets and mifepristone tablets were OOS in 57.1% and 23.7% of samples respectively. With the ongoing global rise in use of medical abortion drugs and increasing availability, together with their role in enabling women to self-manage their abortions, urgent attention is needed to address quality issues [2].

These findings support previous studies of misoprostol quality and demonstrate that a significant problem persists in relation to the quality of misoprostol in LMIC. Given the use of misoprostol alone for medical abortion in many LMIC settings and its other obstetric uses, this is of serious concern.

There is limited historical data on the quality of mifepristone. Our findings indicate that despite being a more stable molecule, quality concerns are also present, with nearly a quarter of the mifepristone tablets showing non-compliance with one or more of the specifications. These findings suggest that increased vigilance is also necessary for mifepristone products in LMIC.

The study identified one falsified misoprostol product, triggering investigations that identified further falsified products in neighboring countries. Falsified Cytotec® has been found previously in LMIC, and these developments suggest that falsification of medical abortion drugs continues to be a serious concern that warrants proactive monitoring by national regulators.

Previous studies have highlighted the importance of primary packaging integrity to manage the degradation of misoprostol, and adherence to controlled storage conditions [12,16]. Only one misoprostol-containing sample was not blistered in Alu/Alu, and while it was not feasible to measure storage temperatures, we did not observe higher rates of OOS misoprostol tablets collected from sites without controlled storage conditions. Similarly, there was no correlation between the age of misoprostol samples when tested and the rate of OOS, in contrast to findings reported in 2011 to 2016. Findings from this study therefore may indicate that manufacturing processes and compliance with international Good Manufacturing Practice (GMP), both for finished products and API, could be the primary current contributor to misoprostol product quality concerns.

These concerns around manufacturing quality emphasize the importance of stronger regulatory standards both in countries where these products are marketed and in countries of manufacture. Whilst global activities aimed at strengthening regulatory capacity in LMIC are ongoing, in the short-term, countries and manufacturers should more actively make use of existing registration reliance mechanisms such as the WHO Collaborative Registration Procedure to increase the availability of quality-assured mifepristone and misoprostol at country level. National authorities should be encouraged to increase surveillance of medical abortion drugs in their markets and take regulatory actions as appropriate.

Manufacturers of internationally distributed misoprostol products should review their specifications to ensure they are in line with, or as stringent as, the Ph. Int. specifications. Similarly, national regulatory authorities should require this for any new misoprostol submissions. For mifepristone, the development of a pharmacopeial monograph for finished product and API would support resolving some of the issues identified within the study.

Prequalification by WHO or an approval from an SRA/ WHO Listed Authority (WLA) [25] provides reliable assurance of product quality. Most UN agencies, USAID, and most European government donors providing funding for abortion related activities have adopted this standard as the minimum requirement in their procurement policies for misoprostol and mifepristone products. However, this standard is not fully harmonized across all funders, nor uniformly reflected in the procurement policies of all SMOs and other organizations engaged in the purchase and supply of medical abortion drugs. In this study, samples which met these standards recorded robust results with a non-compliance rate of 9.1% (one of eleven), compared with 63.5% for products not meeting this standard, with the single quality-assured sample OOS representing a very minor excursion. This finding provides a compelling argument for increased harmonization of procurement policies. Furthermore, the limited number of medical abortion products meeting quality assurance criteria and lack of widespread availability in LMIC indicate a need to increase the number of medical abortion drugs prequalified by WHO, as well as ensuring access to them through obtaining regulatory approval at country level.

A significant number of products available in LMIC are provided by and through SMOs, who are responsible for ensuring their products are quality-assured. Of the 15 tested samples marketed by SMOs, four (26.7%) were observed to be non-compliant with specifications, compared to 63.3% for the non-SMO products. While SMO-marketed products had a lower rate of non-compliance compared to the overall figures, the findings suggest that further work is needed to ensure quality standards are met.

Our analysis did not find any correlation between purchase price and rates of OOS samples tested across the range of product types. The perceived high cost of quality-assured medical abortion drugs as a barrier to their purchase and supply to LMIC should be investigated further. At the present time, there is a lack of incentive for manufacturers to achieve WHO prequalification based on the knowledge that the LMIC marketplace is primarily populated by products not meeting robust quality criteria, which is believed to be reflected in their pricing. A multi-country analysis to correlate LMIC market prices with cost of goods in order establish detailed pricing structures could be undertaken to identify solutions and encourage manufacturers to achieve quality-assured status of their products.

WHO recommends self-managed medical abortion up to 12 weeks’ gestation where there is access to accurate information and a healthcare provider if needed or wanted [2]. In practice, emerging evidence is indicating that access to and self-administration of medical abortion drugs is increasing, with women purchasing from pharmacists, drug sellers and by direct mail from the internet. As more women and girls are empowered through self-care, it will become even more critical that they are able to access quality-assured drugs to ensure the safest outcomes when undertaken without clinical supervision [4].

### Limitations

This study was not designed to provide accurate estimates of the prevalence of poor-quality medicines in each country: the non-random selection of sampling sites and products; the use of the overt sampling method; and the limited sample size in each individual country, all limit such an extrapolation.

Due to sample size limitations, we were not able to assess other quality risks for the products, such as microbial contamination, dissolution for misoprostol, uniformity of mass for mifepristone and water content.

All samples were tested for each tablet type according to the same specification to enable comparison of samples from different manufacturers. International reference pharmacopoeia specifications were used to the maximum extent possible. However, products marketed in LMIC are manufactured to a wide range of manufacturer specifications and methods, and these specifications and methods may be approved by regulatory authorities in individual countries.

We cannot exclude problems with storage and transportation conditions since we sampled only at the point of sale. However, our analysis of basic storage conditions suggested no correlation to OOS findings.

Finally, product testing only provides a snapshot of a small sample of each batch of product, tested at a particular point in its shelf-life. It is a more reliable indicator of product quality when manufacturing quality and storage conditions are known to be acceptable. However, the study does flag important issues and concerns which warrant more detailed investigation, both globally and in the relevant countries.

## Conclusions

Misoprostol products in several geographically diverse, large countries have significant quality issues. Since primary packaging has improved, it leaves API and FPP manufacturing as the probable main causes of substandard misoprostol products.

Although smaller in comparison, this study also provides evidence of mifepristone quality issues with 23.7% of samples tested non-compliant.

Self-administration of medical abortion drugs presents an urgent additional dynamic in the quality debate and evidence-based quality assurance of mifepristone and misoprostol must be a prerequisite for product supply.

## Declarations

### Author Contributions

LC conceived and conceptualized the study, developed abstract, discussion, and conclusions and reviewed. JB developed study protocol, coordinated sampling and testing activities, developed methods, results and limitations, and provided editorial contributions throughout. WI and AT were expert technical leads for the study and established, transferred and validated testing methods. CK led sample collection and coordination with country resources and provided editorial contributions. PP was the operational Manager for the duration of the study. AF coordinated study activities and manuscript development. AMG provided expert guidance throughout the study and was lead reviewer. All authors have read, reviewed and approved the manuscript.

## Data Availability

The data that support the findings of this study are available on request from the corresponding author. The data are not publicly available due to privacy or ethical reasons.

## List of abbreviations

API: Active pharmaceutical ingredient
CRS: Chemical reference standard
EDQM: European Directorate for the Quality of Medicines
EML: Essential Medicines List
FDA: Food and Drug Authority
FPP: Finished pharmaceutical product
GMP: Good Manufacturing Practice
HCI: Health Concepts International
HPLC: High-performance liquid chromatography
ICH: International Council for Harmonization of Technical Requirements for Pharmaceuticals for Human Use
InphA: Institute for Pharmaceutical and Applied Analytics
IPPF: International Planned Parenthood Federation
ISF: Incidents and Substandard / Falsified Medical Products
LMIC: Lower– and middle-income countries
LOQ: Limit of quantitation
OOS: Out of specification
Ph. Int.: International Pharmacopoeia
PQP: Prequalification Program
QA: Quality assurance
SCF: Sample collection form
SMO: Social marketing organization
SOP: Standard operating procedure
SRA: Stringent regulatory authority
UK: United Kingdom
UN: United Nations
USAID: United States Agency for International Development
USD: United States Dollar
USP: United States Pharmacopoeia
WHO: World Health Organization
WLA: WHO-Listed Authority

## Acknowledgments

Peter Hall, China Resources Zizhu Pharmaceuticals Co. Ltd., China and Everlight Chemical Industrial Corporation, Taiwan who provided impurity reference standards, and Nhomsai Hagen who helped with specific questions.

## Figures and Tables

## Additional files

**Additional File 1, .doc file, Tables S1-S8**: specifications for misoprostol and mifepristone tablets for each test conducted, analytical method verification and sample analysis particulars, number of samples collected by site and sampling method, product-specific sample data (i.e. labelled storage requirements, packaging type, number of manufacturers), shelf-life of tested samples, full summary of misoprostol results, and full summary of mifepristone results.

**Additional File 2, .jpg file, Fig.S1**: graph of misoprostol related substances results (full scale).

**Additional File 3, .jpg file, Fig.S2**: graph of mifepristone assay results for each sample.

**Additional File 4, .jpg file, Fig.S3**: rate of OOS results for products sponsored by type of organization.

