## Supplemental Tables for "Quality testing of mifepristone and misoprostol in 11 countries"

Additional File 1: Supplementary Tables

**Table S1: Specifications for misoprostol tablets**

| **TEST** | **ACCEPTANCE CRITERIA** | **ANALYTICAL METHOD** |
| --- | --- | --- |
| Identification (by HPLC) | Misoprostol is confirmed by HPLC chromatogram | HPLC, Ph. Int. |
| Assay* | 90.0 – 110.0% | HPLC, Ph. Int. |
| Uniformity of Content | For n=10, each single unit contains within ±15% of the average amount of the active ingredient. For n=30, no more than one out of 30 units deviates by more than ±15% of the average amount, and none by more than ±25% | HPLC, Ph. Int. |
| Related Substances** | a. Sum of Impurity A, B and E ≤3.0% b. Impurity C ≤1.5% c. Impurity D ≤1.0% | HPLC, Ph. Int. |

** Use the average of the 10 individual results obtained in the test for “Uniformity of content”.*

****	*Impurity A = 8-epi-misoprostol; Impurity B =* *12-epi-misoprostol; Impurity C = misoprostol A; Impurity D =* *misoprostol B; Impurity E = Methyl rac-(13E,16RS)-11α,16-dihydroxy-16,18-dimethyl-9-oxo-20-norprosta-13,17-dien-1-oate (mixture of 4 stereoisomers)*

**Table S2: Specifications for mifepristone tablets**

| **TEST** | **ACCEPTANCE CRITERIA** | **ANALYTICAL METHOD** |
| --- | --- | --- |
| Identification (by HPLC) | Mifepristone is confirmed by HPLC chromatogram | HPLC,  in-house* |
| Dissolution (by USP Apparatus II at 75 rpm, in 900mL of 0.01 N HCl)** | Not less than 80% (Q) of the labelled amount of mifepristone is dissolved in 30 minutes. | HPLC,  in-house |
| Assay | 90.0 – 110.0% | HPLC, in-house* |
| Related Substances*** | a. Demethylated derivative† ≤0.5% b. Individual unknown impurity ≤0.2% c. Total impurities ≤1.0% | HPLC,  in-house* |

***	*Adapted from the related substances method in the USP pending monograph of mifepristone API, 2008*

****	*Based on FDA-recommended dissolution method for mifepristone tablet*

**** The limits are based on the USP pending monograph of mifepristone API, 2008, except for any individual unknown impurity which was established in line with the identification threshold provided in the ICH Q3B guideline (impurities in new drug products)*

*† 11b-[p-(Methylamino)phenyl]-17b*

**Table S3: Analytical method verification and sample analysis particulars**

|  | **Misoprostol** | **Mifepristone** |
| --- | --- | --- |
| Reference products | Cytotec 200 mcg tablets (Lot no. B21291, Pfizer), obtained by InphA from the German market | Mifegyne 200 mg tablets (Lot no. 216, Nordic Pharma), obtained by InphA from the German market |
| Chemical reference standards | Misoprostol chemical reference standard (CRS) (batch no. 3.0) was procured from European Directorate for the Quality of Medicines (EDQM). Reference standards for misoprostol impurities were provided by Everlight Chemical Industrial Corporation, Taiwan | Mifepristone USP-RS (Lot no. F0H385) was obtained from Sigma Aldrich, and demethylated mifepristone was supplied by China Resources Zizhu Pharmaceuticals Co. Ltd, China |
| Analytical methods | 1. HPLC method for identification and content uniformity/assay   - Based on Ph. Int. monograph for misoprostol tablets - 10 tablets were analysed independently, and individual results reported for content uniformity and average results reported for assay | 1. In-house HPLC method for identification, assay and determination of related substances   - Based on the USP pending monograph of mifepristone API, 2008 - Agilent HPLC system; SUPLEX PKB-100 column; 5 μm, 250 x 4.6 mm (Supelco); 0.05 M Phosphate solution/acetonitrile (55/45 %v/v) as mobile phase; flow rate of 1.2 ml/min; injection volume of 10 μl, and; wavelength of 250 nm (UV detection) - 3 tablets were analysed independently and average results reported |
|  | 2. HPLC method for determination of related substances   - Based on Ph. Int. monograph for misoprostol tablets - 5 pooled tablets were analysed, instead of 10 tablets as described in the Ph. Int. monograph, due to limited samples | 2. In-house dissolution method   - Based on the FDA-recommended dissolution method for mifepristone tablet - Paddle apparatus (SOTAX AT Xtend) at 75 rpm, with 900 ml of 0.01 N HCl as dissolution medium. Samples were drawn after 30 minutes through an in-line filter and assayed using the above HPLC method - 6 tablets were analysed for stage 1; if failed, then 6 additional tablets were analysed for stage 2 |
| Method verification | Specificity, repeatability, linearity, accuracy  and limit of quantitation (for related substances) of the methods were successfully verified | Specificity, repeatability, linearity, accuracy  and limit of quantitation (for related substances) of the methods were successfully verified* |

** Method verification was performed as this in-house method was previously fully validated by Health Concepts International (HCI) Ltd in Thailand, a subsidiary of Concept Foundation and a WHO-prequalified quality control laboratory until its closure in 2018*

**Table S4: Number of samples collected, by site and sampling method**

| **Type of Collection Site** | **# Samples** |
| --- | --- |
| 1) Pharmacy attached to a public hospital or clinic | 3 |
| 2) Pharmacy attached to a private hospital or clinic | 6 |
| 3) Pharmacy attached to an NGO clinic | 4 |
| 4) Private pharmacy (stand-alone/retail) | 48 |
| 5) Drug shop or seller | 9 |
| 6) Other* | 24 |
| **Qualification of seller** |  |
| A) Pharmacist | 39 |
| B) Pharmacy worker | 27 |
| C) Drug Seller | 7 |
| D) Other health worker | 8 |
| E) Unknown | 13 |
| **Approach used** |  |
| 1) Mystery shopper | 31 |
| 2) Overt sampling | 57 |
| 3) Hybrid approach | 6 |
| **Observed storage conditions at site** |  |
| A) Enclosed indoor dispensary, air-conditioned or similar | 59 |
| B) Enclosed indoor dispensary, fans | 28 |
| C) Open air premises | 7 |

** Primarily were distributors or sub-outlets (20), but also included two samples provided directly at clinics, one at a central market, and one from an undefined seller.*

**Table S5: Product-specific sample data**

| **Labelled storage requirements for temperature** | **Combipack products** | **Misoprostol-only products** | **Mifepristone-only products** | **All samples** |
| --- | --- | --- | --- | --- |
| Not stated* | 13 | 4 | 1 | 18 |
| Under 25°C | 11 | 6 | 0 | 17 |
| Under 30°C | 20 | 21 | 8 | 49 |
| **Primary packaging type** |  |  |  |  |
| Alu/Alu blister | 41 | 30 | 2 | 73 |
| Plastic/Alu blister | 0 | 1 | 7 | 8 |
| Sealed in foil pouch** | 3 | 0 | 0 | 3 |
| **Manufacturers** |  |  |  |  |
| # of manufacturers |  |  |  | 42 |
| # of countries of manufacture |  |  |  | 10 |
| **Total batch samples** | **44** | **31** | **9** | **84** |

** Storage temperature requirement was non-specific. These samples were primarily labelled to store in a cool, dry, dark place / protect from light.*

*** 3 combipack samples from Bangladesh were packed in a foil over the primary blister packaging. For one, the package insert reported the primary packaging as being alu/alu; whereas the package insert of the other two samples, different batches of the same product, did not report the packaging material.*

**Table S6: Shelf-life of samples tested per product type**

| **Shelf-life** | **Combipack products** | **Misoprostol-only products** | **Mifepristone-only products** | **All products** |
| --- | --- | --- | --- | --- |
| 12 months | 0 | 1 | 0 | **1** |
| 18 months | 2 | 0 | 0 | **2** |
| 24 months | 27 | 13 | 2 | **42** |
| 30 months | 1 | 0 | 0 | **1** |
| 36 months | 1 | 11 | 3 | **15** |
| 48 months | 0 | 0 | 1 | **1** |
| Unknown* | 0 | 1 | 1 | **2** |

***Overall findings***

**Table S7: Summary of misoprostol samples testing**

| **No.** | **Report ID** | **Sampling country** | **Type of product** | **COM** | **Mfg date / Exp date** | **Age of sample at time of analysis (months)** ^#^ | **Packaging** | **ID** | **Mean assay [90.0-110.0%)** | **CU**  **[±15% of the average]** | **Sum of Impurity A, B and E [NMT 3.0%]** | **Impurity C**  **[NMT 1.5%]** | **Impurity D**  **[NMT 1.0%]** |
| --- | --- | --- | --- | --- | --- | --- | --- | --- | --- | --- | --- | --- | --- |
| 1 | P-M-210445 | Burkina Faso | Combipack | India | Feb-20 / Jan-22 | 14 | Alu/Alu blister pack | Positive | 101.0 | Comply | 0.8 | 0.8 | 0.0 |
| 2 | P-M-210446 | Burkina Faso | Misoprostol-only | India | Aug-20 / Jul-22 | 8 | Alu/Alu blister pack | Positive | 100.6 | Comply | 0.7 | 0.4 | 0.0 |
| 3 | P-M-210184 | Cambodia | Combipack | India | Sep-19 / Aug-21 | 17 | Alu/Alu blister pack | Positive | 90.7 | Comply | 0.8 | 1.9 | 0.1 |
| 4 | P-M-210185 | Cambodia | Combipack | India | Aug-19 / Jul-21 | 19 | Alu/Alu blister pack | positive | 100.1 | Comply | 0.7 | 0.7 | 0.0 |
| 5 | P-M-210186 | Cambodia | Combipack | India | Sep-19 / Aug-21 | 17 | Alu/Alu blister pack | positive | 104.4 | Comply | 0.8 | 0.4 | 0.2 |
| 6 | P-M-210187 | Cambodia | Combipack | India | Sep-19 / Aug-21 | 18 | Alu/Alu blister pack | positive | 96.6 | Comply | 1.3 | 6.9 | 0.4 |
| 7 | P-M-210188 | Cambodia | Combipack | India | Dec-19 / Nov-21 | 15 | Alu/Alu blister pack | positive | 100.4 | Comply | 0.8 | 0.6 | 0.1 |
| 8 | P-M-210189 | Cambodia | Misoprostol-only | India | Sep-19 / Aug-21 | 18 | Alu/Alu blister pack | positive | 97.9 | Comply | 1.4 | 3.6 | 0.4 |
| 9 | P-M-210466 | DRC | Misoprostol-only | India | Aug-19 / Jul-21 | 22 | Alu/Alu blister pack | positive | 100.6 | comply | 1.1 | 1.9 | 0.1 |
| 10 | P-M-210467 | DRC | Misoprostol-only | India | Sep-20 / Aug-22 | 8 | Alu/Alu blister pack | positive | 98.4 | comply | 5.5 | 0.9 | 0.0 |
| 11 | P-M-210468 | DRC | Misoprostol-only | India | Jun-20 / May-22 | 11 | Alu/Alu blister pack | positive | 86.7 | comply | 5.3 | 2.3 | 0.1 |
| 12 | P-M-210469 | DRC | Misoprostol-only | India | Jun-20 / May-22 | 11 | Alu/Alu blister pack | positive | 96.6 | comply | 0.7 | 0.5 | 0.0 |
| 13 | P-M-210470 | DRC | Misoprostol-only | UK | May-19 / May-22 | 24 | Alu/Alu blister pack | negative | X | X | X | X | X |
| 14 | P-M-210471 | DRC | Misoprostol-only | India | Aug-20 / Jul-23 | 9 | Alu/Alu blister pack | positive | 35.7 | not comply (Stage 1)* | 22.2 | 196.4 | 8.3 |
| 15 | P-M-210655 | India | Combipack | India | Nov-20 / Oct-22 | 10 | Alu/Alu blister pack | positive | 102.1 | comply | 0.9 | 1.7 | 0.0 |
| 16 | P-M-210912 | India | Combipack | India | Jul-20 / Jun-22 | 14 | Alu/Alu blister pack | positive | 91.5 | comply | 5.2 | 2.0 | 0.1 |
| 17 | P-M-210913 | India | Combipack | India | Mar-21 / Feb-23 | 8 | Alu/Alu blister pack | positive | 109.0 | comply (at Stage 2) | 1.0 | 1.1 | 0.1 |
| 18 | P-M-210914 | India | Combipack | India | Jul-20 / Jun-22 | 14 | Alu/Alu blister pack | positive | 90.8 | comply | 3.5 | 1.4 | 0.9 |
| 19 | P-M-210915 | India | Combipack | India | Feb-21 / Jul-22 | 7 | Alu/Alu blister pack | positive | 113.7 | comply | 5.3 | 1.2 | 0.2 |
| 20 | P-M-210975 | India | Combipack | India | Aug-20 / Jul-22 | 15 | Alu/Alu blister pack | positive | 100.7 | comply | 0.8 | 1.0 | 0.1 |
| 21 | P-M-210976 | India | Combipack | India | Jul-20 / Jun-22 | 16 | Alu/Alu blister pack | positive | 93.3 | comply | 6.6 | 1.5 | 0.6 |
| 22 | P-M-210977 | India | Combipack | India | Apr-21 / Mar-23 | 7 | Alu/Alu blister pack | positive | 86.2 | comply | 3.9 | 3.2 | 26.8 |
| 23 | P-M-210978 | India | Combipack | India | Feb-21 / Jan-23 | 9 | Alu/Alu blister pack | positive | 75.3 | not comply (Stage 1)* | 16.8 | 17.5 | 12.3 |
| 24 | P-M-210979 | India | Combipack | India | Oct-20 / Sep-22 | 13 | Alu/Alu blister pack | positive | 86.9 | comply | 4.7 | 2.6 | 0.6 |
| 25 | P-M-211028 | India | Combipack | India | Jan-21 / Dec-22 | 10 | Alu/Alu blister pack | positive | 88.6 | comply | 4.5 | 2.0 | 0.1 |
| 26 | P-M-211029 | India | Combipack | India | Dec-20 / Nov-22 | 11 | Alu/Alu blister pack | positive | 92.1 | comply | 3.2 | 1.5 | 0.3 |
| 27 | P-M-211030 | India | Combipack | India | Mar-21 / Feb-23 | 8 | Alu/Alu blister pack | positive | 99.3 | comply | 0.8 | 1.3 | 1.9 |
| 28 | P-M-211031 | India | Combipack | India | Jun-20 / May-22 | 17 | Alu/Alu blister pack | positive | 93.4 | comply | 4.2 | 1.4 | 0.4 |
| 29 | P-M-210455 | Kyrgyzstan | Misoprostol-only | Russia | n.s. / Sep-22 | unknown | Plastic/Alu blister pack | positive | 100.8 | comply | 0.8 | 2.3 | 0.1 |
| 30 | P-M-210456 | Kyrgyzstan | Misoprostol-only | China | Jun-19 / May-21 | 23 | Alu/Alu blister pack | positive | 103.4 | comply | 0.1 | 0.6 | 0.0 |
| 31 | P-M-210453 | Moldova | Combipack | India | Aug-20 / Jul-22 | 8 | Alu/Alu blister pack | positive | 103.1 | comply | 0.4 | 0.4 | 0.0 |
| 32 | P-M-210459 | Nepal | Combipack | India | Oct-20 / Sep-22 | 7 | Alu/Alu blister pack | positive | 101.9 | comply | 0.5 | 0.2 | 0.0 |
| 33 | P-M-210460 | Nepal | Combipack | Nepal | Dec-20 / Nov-22 | 6 | Alu/Alu blister pack | positive | 101.9 | comply | 0.8 | 0.6 | 0.0 |
| 34 | P-M-210461 | Nepal | Combipack | Nepal | Oct-20 / Sep-22 | 7 | Alu/Alu blister pack | positive | 96.7 | comply | 0.5 | 0.4 | 0.0 |
| 35 | P-M-210462 | Nepal | Combipack | India | May-20 / Oct-21 | 12 | Alu/Alu blister pack | positive | 100.8 | comply | 0.7 | 0.4 | 0.0 |
| 36 | P-M-210463 | Nepal | Combipack | India | Aug-20 / Jul-22 | 9 | Alu/Alu blister pack | positive | 92.4 | comply | 2.0 | 0.5 | 0.0 |
| 37 | P-M-210465 | Nepal | Misoprostol-only | Nepal | Jan-21 / Dec-22 | 5 | Alu/Alu blister pack | positive | 100.3 | comply | 0.8 | 0.3 | 0.0 |
| 38 | P-M-210518 | Nigeria | Misoprostol-only | India | Jul-19 / Jun-22 | 22 | Alu/Alu blister pack | positive | 85.4 | comply | 7.4 | 11.7 | 1.3 |
| 39 | P-M-210519 | Nigeria | Misoprostol-only | India | Dec-19 / Nov-22 | 18 | Alu/Alu blister pack | positive | 114.3 | comply | 5.6 | 2.5 | 0.1 |
| 40 | P-M-210520 | Nigeria | Misoprostol-only | India | Dec-19 / Nov-22 | 18 | Alu/Alu blister pack | positive | 77.2 | comply | 8.7 | 29.5 | 5.5 |
| 41 | P-M-210522 | Nigeria | Combipack | India | Mar-20 / Feb-23 | 15 | Alu/Alu blister pack | positive | 13.4 | not comply (Stage 1)* | 15.9 | 84.5 | 95.1 |
| 42 | P-M-210523 | Nigeria | Combipack | India | Oct-20 / Mar-23 | 8 | Alu/Alu blister pack | positive | 28.4 | not comply (stage 1)* | 19.0 | 52.3 | 76.1 |
| 43 | P-M-210524 | Nigeria | Misoprostol-only | Nigeria | Nov-20 / Nov-22 | 7 | Alu/Alu blister pack | positive | 104.5 | comply | 0.5 | 0.7 | 0.0 |
| 44 | P-M-210525 | Nigeria | Misoprostol-only | UK | Feb-20 / Jan-23 | 16 | Alu/Alu blister pack | positive | 100.7 | comply | 0.7 | 0.4 | 0.1 |
| 45 | P-M-201163 | Pakistan | Misoprostol-only | Pakistan | Oct-20 / Sep-22 | 8 | Alu/Alu blister pack | positive | 104.2 | comply | 0.8 | 0.9 | 0.0 |
| 46 | P-M-201164 | Pakistan | Misoprostol-only | Pakistan | Oct-20 / Oct-21 | 8 | Alu/Alu blister pack | positive | 96.9 | comply | 2.6 | 13.0 | 0.6 |
| 47 | P-M-210262 | Pakistan | Misoprostol-only | Pakistan | Sep-20 / Sep-22 | 9 | Alu/Alu blister pack | positive | 82.9 | comply | 1.4 | 1.9 | 0.1 |
| 48 | P-M-210447 | Uganda | Misoprostol-only | UK | Feb-20 / Jan-23 | 14 | Alu/Alu blister pack | positive | 101.3 | comply | 0.7 | 0.4 | 0.0 |
| 49 | P-M-210448 | Uganda | Misoprostol-only | India | Jul-20 / Jun-23 | 9 | Alu/Alu blister pack | positive | 97.6 | comply | 0.7 | 1.1 | 0.0 |
| 50 | P-M-210449 | Uganda | Misoprostol-only | India | Jan-20 / Dec-21 | 15 | Alu/Alu blister pack | positive | 96.4 | comply | 1.3 | 2.0 | 0.0 |
| 51 | P-M-210450 | Uganda | Combipack | India | Jun-20 / May-22 | 10 | Alu/Alu blister pack | positive | 96.7 | comply | 0.8 | 0.6 | 0.1 |
| 52 | P-M-210452 | Uganda | Combipack | India | May-20 / Apr-22 | 11 | Alu/Alu blister pack | positive | 101.8 | comply | 0.6 | 0.3 | 0.0 |
| 53 | P-M-210533 | Vietnam | Misoprostol-only | Vietnam | 17/12/2019 / 17/12/2022 | 18 | Alu/Alu blister pack | positive | 98.7 | comply | 1.3 | 3.6 | 0.2 |
| 54 | P-M-210588 | Vietnam | Misoprostol-only | Korea | 27/12/2019 / 26/12/2022 | 18 | Alu/Alu blister pack | positive | 100.4 | comply | 0.6 | 1.9 | 0.1 |
| 55 | P-M-210731 | Vietnam | Misoprostol-only | Vietnam | 23/03/2020 / 22/03/2023 | 18 | Alu/Alu blister pack | positive | 106.9 | comply | 1.7 | 8.3 | 0.4 |
| 56 | P-M-210748 | Vietnam | Misoprostol-only | Vietnam | 05/05/2021 / 04/05/2023 | 6 | Alu/Alu blister pack | positive | 105.1 | comply | 0.3 | 0.8 | 0.0 |

COM = Country of manufacture; ID = Identification; NMT = Not more than; n.s. = not specified; X = not performed

Analytical results which are out of specification are highlighted in red.

^#^ Age of sample was based on the month misoprostol assay was tested vs the month of manufacture

* Stage 2 content uniformity testing considered unnecessary due to assay out-of-specification

**Table S8: Summary of mifepristone samples testing**

| **No.** | **Report ID** | **Sampling country** | **Type of product** | **Country of manufacture** | **Mfg date / Exp date** | **Age of sample at time of analysis (months)^#^** | **Packaging** | **ID** | **Mean assay (RSD)**  **[90.0-110.0%]** | **Demethylated impurity**  **[NMT 0.5%]** | **Any single impurities [NMT 0.2%]** | **Total impurities [NMT 1.0%]** | **Mean dissolution**  **(range)**  **[Q=85% in 30 mins]** |
| --- | --- | --- | --- | --- | --- | --- | --- | --- | --- | --- | --- | --- | --- |
| 1 | P-M-210445 | Burkina Faso | Combipack | India | Feb-20 / Jan-22 | 14 | Alu/Alu blister pack | positive | 101.5  (0.4) | 0.3 | comply | 0.4 | 91  (90 - 94) |
| 2 | P-M-210184 | Cambodia | Combipack | India | Sep-19 / Aug-21 | 21 | Alu/Alu blister pack | positive | 96.2  (1.3) | 0.4 | comply | 0.9 | 104  (100 - 106) |
| 3 | P-M-210185 | Cambodia | Combipack | India | Aug-19 / Jul-21 | 23 | Alu/Alu blister pack | positive | 98.1  (0.1) | 0.5 | comply | 0.7 | 100  (99 - 101) |
| 4 | P-M-210186 | Cambodia | Combipack | India | Sep-19 / Aug-21 | 21 | Alu/Alu blister pack | positive | 99.7  (0.2) | 0.4 | comply | 0.5 | 101  (97 - 103) |
| 5 | P-M-210187 | Cambodia | Combipack | India | Sep-19 / Aug-21 | 21 | Alu/Alu blister pack | positive | 92.5  (1.2) | 0.5 | comply | 0.6 | 100  (99 - 101) |
| 6 | P-M-210188 | Cambodia | Combipack | India | Dec-19 / Nov-21 | 18 | Alu/Alu blister pack | positive | 102.0  (N/A)* | 0.3 | comply | 0.4 | X |
| 7 | P-M-210655 | India | Combipack | India | Nov-20 / Oct-22 | 10 | Alu/Alu blister pack | positive | 97.9  (0.7) | 0.3 | comply | 0.5 | 94  (94 - 95) |
| 8 | P-M-210912 | India | Combipack | India | Jul-20 / Jun-22 | 14 | Alu/Alu blister pack | positive | 96.8  (0.9) | 0.4 | not comply | 0.9 | 93  (91 - 94) |
| 9 | P-M-210913 | India | Combipack | India | Mar-21 / Feb-23 | 6 | Alu/Alu blister pack | positive | 97.1  (1.0) | 0.4 | comply | 0.5 | 98  (97 - 100) |
| 10 | P-M-210914 | India | Combipack | India | Jul-20 / Jun-22 | 14 | Alu/Alu blister pack | positive | 95.4  (0.6) | 0.4 | comply | 0.5 | 95  (94 - 96) |
| 11 | P-M-210915 | India | Combipack | India | Feb-21 / Jul-22 | 7 | Alu/Alu blister pack | positive | 92.9  (0.3) | 0.2 | comply | 0.3 | 89  (86 - 96) |
| 12 | P-M-210975 | India | Combipack | India | Aug-20 / Jul-22 | 15 | Alu/Alu blister pack | positive | 97.5  (0.9) | 0.5 | comply | 0.7 | 96  (93 - 97) |
| 13 | P-M-210976 | India | Combipack | India | Jul-20 / Jun-22 | 16 | Alu/Alu blister pack | positive | 95.6  (0.4) | 0.5 | not comply | 1.2 | 97  (96 - 98) |
| 14 | P-M-210977 | India | Combipack | India | Apr-21 / Mar-23 | 7 | Alu/Alu blister pack | positive | 91.1  (3.4) | 0.2 | comply | 0.2 | 93  (92 - 95) |
| 15 | P-M-210978 | India | Combipack | India | Feb-21 / Jan-23 | 9 | Alu/Alu blister pack | positive | 84.5  (0.4) | 0.2 | not comply | 2.6 | 77  (75 - 80)** |
| 16 | P-M-210979 | India | Combipack | India | Oct-20 / Sep-22 | 13 | Alu/Alu blister pack | positive | 95.5  (1.0) | 0.3 | comply | 0.4 | 97  (96 - 98) |
| 17 | P-M-211028 | India | Combipack | India | Jan-21 / Dec-22 | 10 | Alu/Alu blister pack | positive | 95.2  (1.7) | 0.3 | comply | 0.4 | 88  (88 - 89) |
| 18 | P-M-211029 | India | Combipack | India | Dec-20 / Nov-22 | 11 | Alu/Alu blister pack | positive | 101.0  (1.3) | 0.2 | not comply | 0.8 | 85  (83 - 86)** |
| 19 | P-M-211030 | India | Combipack | India | Mar-21 / Feb-23 | 8 | Alu/Alu blister pack | positive | 90.2  (4.2) | 0.4 | comply | 0.7 | 87  (83 - 91)** |
| 20 | P-M-211031 | India | Combipack | India | Jun-20 / May-22 | 17 | Alu/Alu blister pack | positive | 97.9  (4.3) | 0.3 | not comply | 0.9 | X |
| 21 | P-M-211061 | India | Combipack | India | May-21 / Apr-23 | 6 | Alu/Alu blister pack | positive | X | X | X | X | 96  (95 - 97) |
| 22 | P-M-210454 | Kyrgyzstan | Mifepristone-alone | Russia | n.s. / Sep-25 | unknown | Plastic/Alu blister pack | positive | 97.4  (0.6) | 0.4 | comply | 0.7 | 99  (98 - 100) |
| 23 | P-M-210453 | Moldova | Combipack | India | Aug-20 / Jul-22 | 8 | Alu/Alu blister pack | positive | 96.9  (0.8) | 0.5 | comply | 0.5 | 96  (93 - 97) |
| 24 | P-M-210459 | Nepal | Combipack | India | Oct-20 / Sep-22 | 7 | Alu/Alu blister pack | positive | 96.7  (3.7) | 0.3 | comply | 0.4 | 95  (93 - 97) |
| 25 | P-M-210460 | Nepal | Combipack | Nepal | Dec-20 / Nov-22 | 6 | Alu/Alu blister pack | positive | 100.2  (0.8) | 0.4 | comply | 0.7 | 96  (93 - 97) |
| 26 | P-M-210461 | Nepal | Combipack | Nepal | Oct-20 / Sep-22 | 7 | Alu/Alu blister pack | positive | 98.8  (0.8) | 0.3 | comply | 0.4 | 96  (94 - 98) |
| 27 | P-M-210462 | Nepal | Combipack | India | May-20 / Oct-21 | 12 | Alu/Alu blister pack | positive | 100.3  (1.5) | 0.3 | comply | 0.4 | 99  (97 - 101) |
| 28 | P-M-210463 | Nepal | Combipack | India | Aug-20 / Jul-22 | 9 | Alu/Alu blister pack | positive | 96.2  (1.0) | 0.3 | comply | 0.6 | 91  (90 - 92) |
| 29 | P-M-210464 | Nepal | Mifepristone-alone | Nepal | Dec-20 / Nov-22 | 6 | Plastic/Alu blister pack | positive | 101.0  (1.0) | 0.3 | comply | 0.4 | 95  (95 - 96) |
| 30 | P-M-210516 | Nigeria | Mifepristone-alone | India | Dec-20 / Nov-23 | 6 | Alu/Alu blister pack | positive | 97.8  (0.3) | 0.2 | comply | 0.6 | 91  (90 - 92) |
| 31 | P-M-210521 | Nigeria | Mifepristone-alone | India | Apr-19 / Mar-22 | 26 | Alu/Alu blister pack | positive | 78.6  (2.0) | 0.4 | not comply | 2.1 | 11  (6 - 16) |
| 32 | P-M-210522 | Nigeria | Combipack | India | Mar-20 / Feb-23 | 15 | Alu/Alu blister pack | positive | 95.2  (1.8) | 0.3 | comply | 0.9 | 83  (80 - 89) |
| 33 | P-M-210523 | Nigeria | Combipack | India | Oct-20 / Mar-23 | 8 | Alu/Alu blister pack | positive | 94.5  (1.2) | 0.2 | comply | 0.5 | 96  (95 - 97) |
| 34 | P-M-210450 | Uganda | Combipack | India | Jun-20 / May-22 | 10 | Alu/Alu blister pack | positive | 95.2 (1.3) | 0.4 | comply | 0.6 | 92  (90 - 93) |
| 35 | P-M-210452 | Uganda | Combipack | India | May-20 / Apr-22 | 11 | Alu/Alu blister pack | positive | 101.5  (1.4) | 0.3 | comply | 0.4 | 98  (88 - 102) |
| 36 | P-M-210534 | Vietnam | Mifepristone-alone | Vietnam | 26/09/2020 / 26/09/2023 | 9 | Plastic/Alu blister pack | positive | 99.8  (0.7) | 0.4 | not comply | 1.0 | 96  (95 - 96) |
| 37 | P-M-210586 | Vietnam | Mifepristone-alone | Vietnam | 25/03/2020 / 24/03/2024 | 15 | Plastic/Alu blister pack | positive | 100.8  (1.8) | 0.5 | comply | 0.8 | 92  (89 - 95) |
| 38 | P-M-210587 | Vietnam | Mifepristone-alone | Vietnam | 17/12/2020 / 16/12/2022 | 6 | Plastic/Alu blister pack | positive | 97.6  (2.4) | 0.5 | comply | 1.0 | 53  (52 - 54) |

COM = Country of manufacture; ID = Identification; NMT = Not more than; Q = Quantity of dissolved active ingredient; n.s. = not specified; X = not performed

Analytical results which are out of specification are highlighted in red.

^#^ Age of sample was based on the month mifepristone assay was tested vs the month of manufacture, except for sample P-M-211061 which was based on when dissolution was tested

* Assay results were based on only 2 single units due to insufficient samples; therefore, RSD calculation was not possible.

** Insufficient samples to proceed with Stage 2 dissolution testing
