## Supplementary figures and images for "Quality testing of mifepristone and misoprostol in 11 countries"

### Supplemental Figure 1

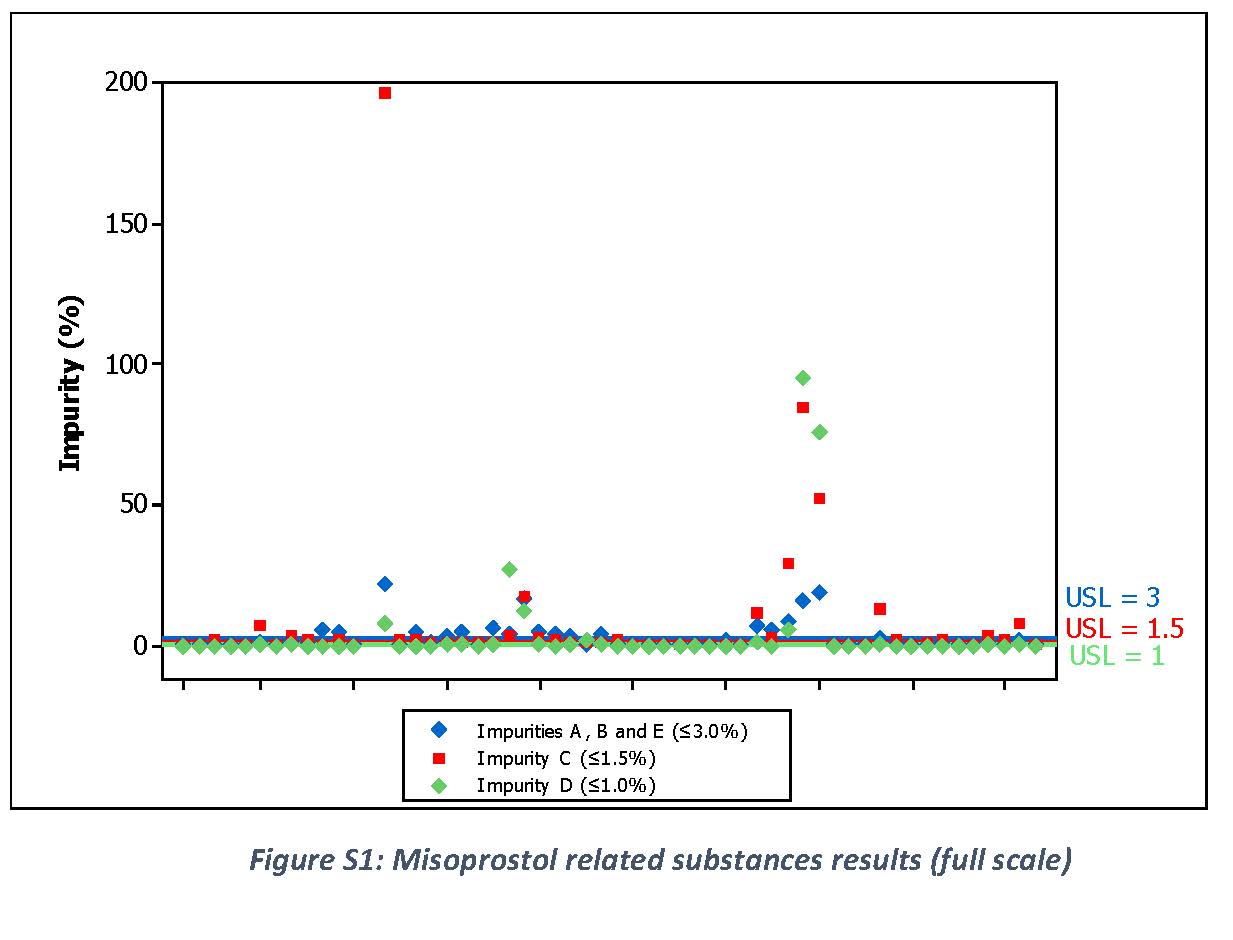

### Supplemental Figure 2

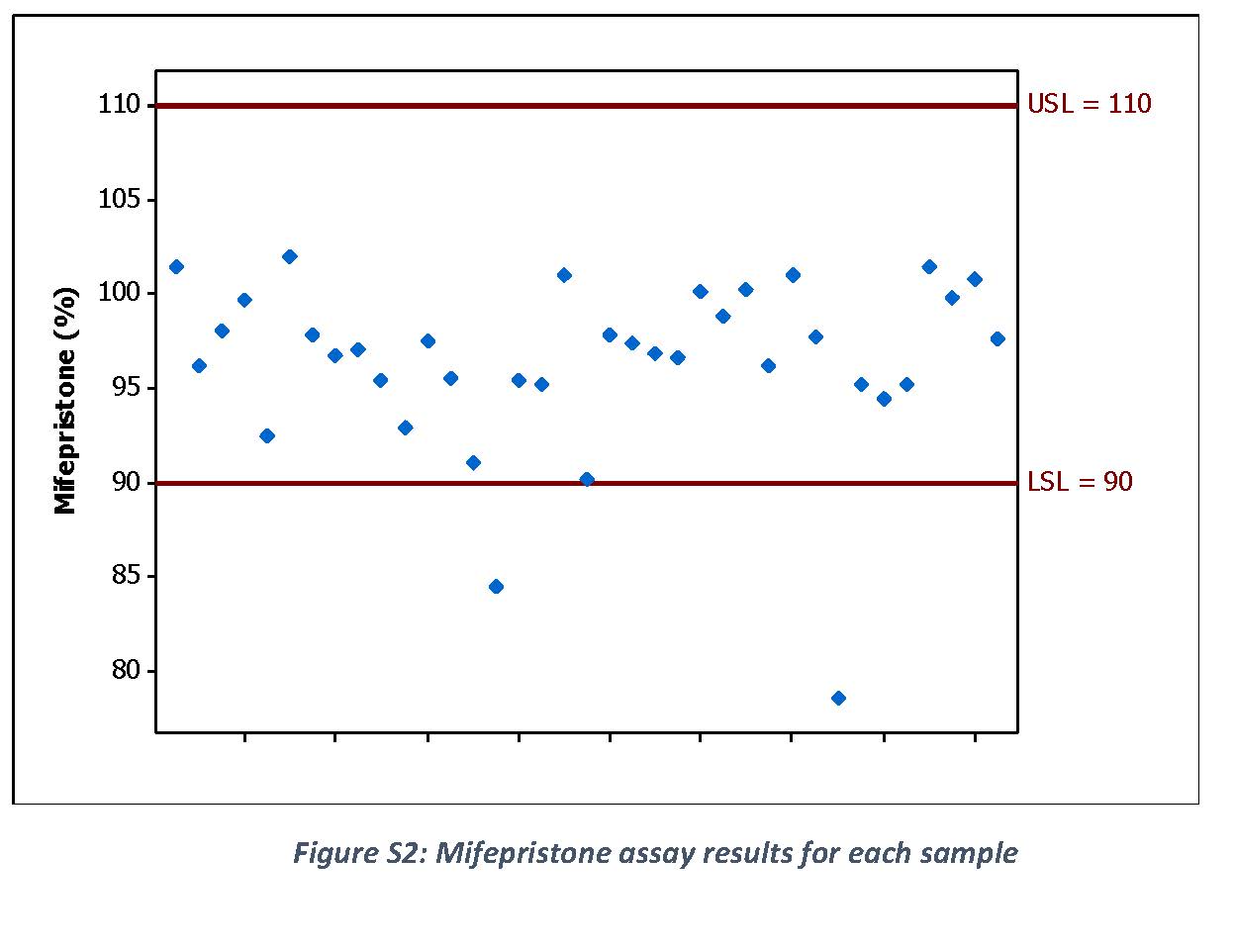

### Supplemental Figure 3

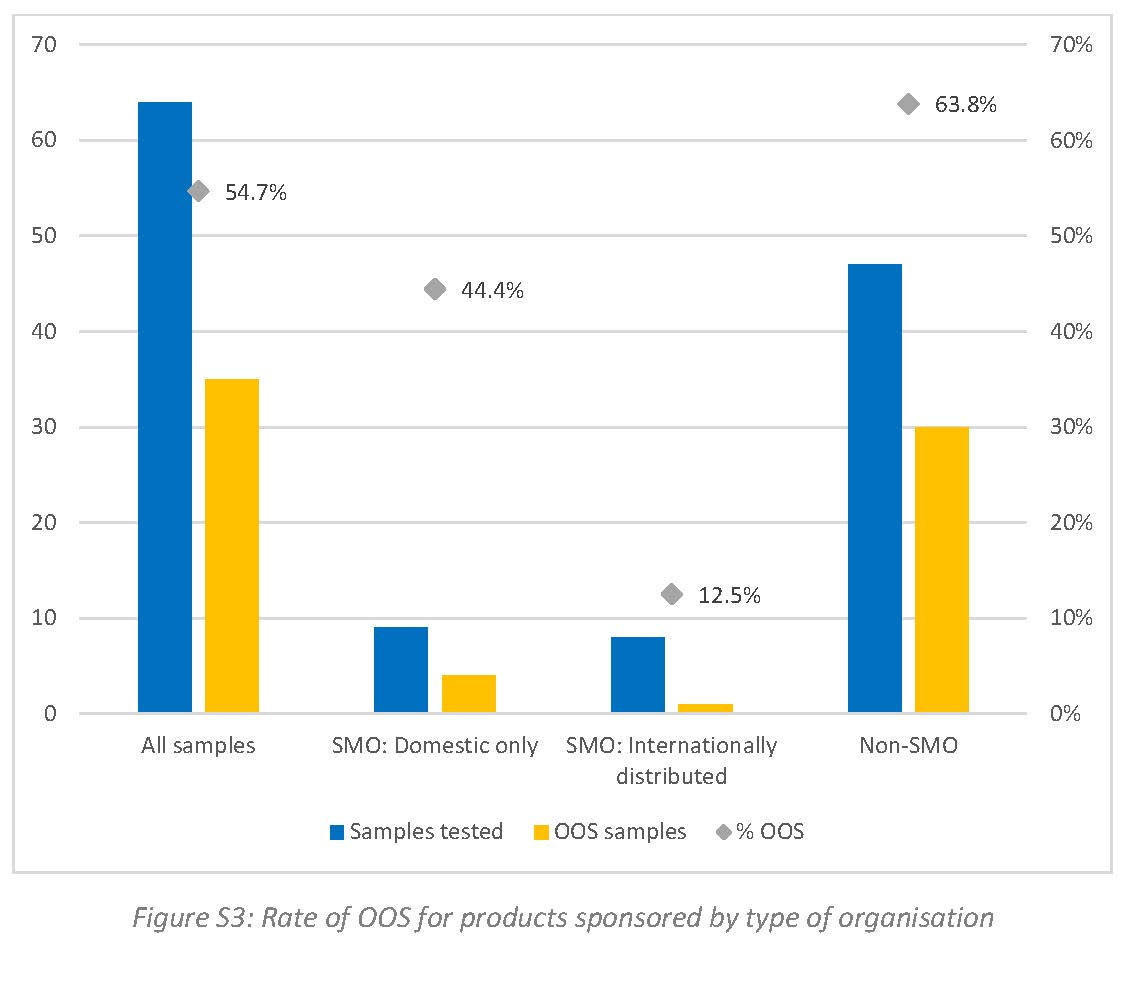
